# Verbal autopsy analysis of childhood deaths in rural Gambia

**DOI:** 10.1101/2022.10.26.22281581

**Authors:** Baleng Mahama Wutor, Isaac Osei, Lobga Babila Galega, Esu Ezeani, Williams Adefila, Ilias Hossain, Golam Sarwar, Grant Mackenzie

## Abstract

**Background:** In low-resource settings, it is challenging to ascertain the burden and causes of under-5 mortality as many deaths occur outside health facilities. Verbal autopsy (VA) is an important tool that provides data on causes of death in communities with limited access to health care. We aimed to determine the causes of childhood deaths by VA in rural Gambia.

**Methodology:** We used WHO standard questionnaires to conduct VAs for deaths under-5 years of age in the Basse and Fuladu West Health and Demographic Surveillance Systems in rural Gambia between September 01, 2019, and December 31, 2021. Two physicians assigned a cause of death and discordant diagnoses were resolved by consensus. Causes of death were classified using the International Classification of Disease 10th edition codes.

**Results:** VAs were conducted for 89% (647/727) of deaths. Of these deaths, 49.5% (n=319) occurred at home, 50.1% (n=324) in females, 37.1% (n=240) in neonates, and 27.1% (n=175) in infants aged 1-11 months. Outside the neonatal period, pneumonia (27.0%, n=110), diarrhoeal diseases (23.3%, n=95), and sepsis (21.6%, n=88) were the commonest primary causes of death. In the neonatal period, unspecified perinatal causes of death (29.6%, n=71), birth asphyxia (23.8%, n=57) and prematurity/low birth weight (17.1%, n=41) were the commonest causes. Severe malnutrition (28.6%, n=185), unspecified perinatal deaths (10.7%, n=69), pneumonia (10.2%, n=66), and prematurity/low birth weight (10.2%, n=66) were the commonest underlying causes of death.

**Conclusion:** According to VA analysis, half of deaths amongst children under-5 in rural Gambia occur at home. Pneumonia, diarrhoea, and sepsis, and the underlying cause of severe malnutrition, as well as birth asphyxia in neonates, remain the predominant causes of child mortality in rural Gambia. Improved health care and health-seeking behaviour may reduce childhood deaths in rural Gambia.

## Introduction

Reliable data on under-5 mortality is needed to effectively track progress towards attaining Sustainable Development Goal 3.2 which seeks to reduce neonatal and under-5 mortality to 12 per 1000 live births and 25 per 1000 live births respectively by 2030[1]. Globally, under-5 mortality in 2019 was estimated to be about 5.2 million[2]. More than half of these deaths occurred in Sub-Saharan Africa. Accurate and complete morbidity and mortality data are essential not only for planning and research purposes but also for measuring the impact of health interventions[3]. This is especially important in low and middle-income countries where resources are limited and need to be prioritized [4].

Although some progress has been made in gathering data for evidence-based decision-making in child health, major gaps still exist[5]. Whereas death registration coverage is nearly 100% in the World Health Organisation (WHO) European Region, coverage is less than 10% in the African Region[6]. Even where Civil Registration and Vital Statistics (CRVS) systems exist in low and middle-income countries, they are often weak and incomplete [7]. Apart from the weak systems in these countries, a significant number of deaths in rural communities occur at home or health posts. Such deaths are not likely to be recorded and consequently, a cause of death will not be assigned. This paucity of mortality data tends to further perpetuate health inequalities[8].

To help fill this gap, the verbal autopsy (VA) was developed and refined over several years to help provide mortality data in countries with weak CRVS systems and in areas where deaths occur at home and are not reported[6, 9, 10]. A VA is a structured questionnaire that is used to ascertain the cause of death by interviewing caregivers or the next of kin who were present during the illness and at the time of death[6]. The caregiver or next of kin is asked about the signs and symptoms the deceased exhibited in the immediate days preceding their demise. Though VAs have several limitations and logistical challenges, they have proved over the years to be a viable option in determining unknown causes of death[11, 12]. This is especially so in sub-Saharan Africa where there are major hurdles to incisional autopsies. Apart from the lack of qualified personnel to perform incisional biopsies, relatives often cite religious and cultural grounds and fears of mutilating the deceased’s body as reasons for refusing such autopsies[13].

Information gathered from a verbal autopsy is used to determine the cause of death using various methods. Some of these methods include Physician-Certified Verbal Autopsy (PCVA)[14] and computer-based probabilistic methods [15, 16]. Some of the computer algorithms that have been developed to assign causes of death using VA data include InterVA4, SmartVA, and InSilicoVA[17-19]. However, the widely used method of VA in sub-Saharan Africa is the PCVA which typically involves two physicians who assign a cause of death based on the information recorded in the VA interview [14, 20]. Even though PCVA can be time-consuming and expensive to undertake, it remains a valuable tool for assigning causes of death for deaths occurring outside the healthcare setting[21]. Given its widespread usage, it has been the most validated type of VA[20].

Where Health and Demographic Surveillance Systems (HDSS) exist, VAs are routinely conducted and provide vital data on mortality at the level of the community. Though HDSS are often difficult and expensive to establish and maintain, they provide a vital platform for research and surveillance activities. There are currently 48 HDSS sites across Africa and Asia which collaborate through the INDEPTH Network to undertake longitudinal collection of health and demographic data in low and middle-income countries[22]. These HDSS sites have been instrumental in filling the gap in capturing mortality data in a range of defined populations[23].

In this study, we sought to characterise causes of death assigned using PCVA within two HDSS sites in The Gambia (Basse and Fuladu West HDSS sites).

## Methodology

### Study Setting

The Gambia is the smallest country in mainland Africa with a population of about 2.4 million. According to the 2018 Gambia Multiple Indicator Cluster Survey, 15.4% of the population are children under-5 years of age and the mortality in this age group is about 57 per 1,000 live births[24]. The Gambia currently does not have a robust CRVS system and many deaths in rural communities go unreported and the causes are not ascertained.

The Medical Research Council Unit The Gambia (MRCG) is a leading biomedical research institution based in The Gambia. Over several decades of its existence, it has established and currently operates four HDSS sites across the country. These are in the Farafenni, West Kiang, Fuladu West, and Basse areas. These platforms not only collect important health and demographic data but also routinely conduct VAs.

The Basse HDSS was established in 2007 in the Upper River Region of the Gambia and has been vital in several field studies[25-27]. It is located in the eastern part of the Gambia on the south bank of the River Gambia and covers an area of approximately 1,100 km^2^. The mostly rural area is inhabited by about 202,000 residents with about 225 villages. Children aged less than 5 years make up 19% of the population. Most of the population practice Islam. The Sarahule and Fula tribes are the commonest ethnic groups in the area. The months from June to October experience a wet season, while the remaining months are dry. Most of the inhabitants are either crop or livestock farmers. The district hospital at Basse is the major health facility in the region and receives referrals from smaller clinics in the area.

The Fuladu West HDSS is located in the Central River Region and extends towards the west along the south bank of the River Gambia. It is made up of 213 villages with a total population of 99,113. The main hospital in the area is Bansang Hospital which receives referrals from the Central River Region as well as the Upper River Region. The Basse and Fuladu West HDSSs currently support the cluster-randomized Pneumococcal Vaccines Schedules clinical trial[28]. Fig 1 below shows a map of the Gambia showing the Basse and Fuladu West HDSS sites.

**Fig 1.**
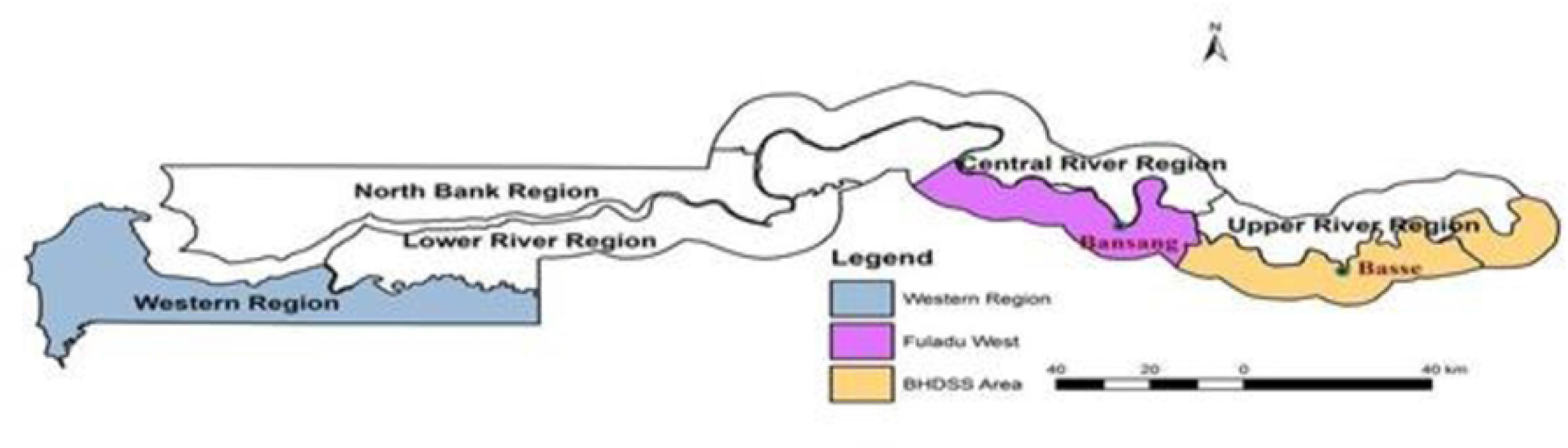
Map of The Gambia highlighting the Basse and Fuladu West HDSS sites.

The Basse District Hospital and Bansang Hospital are the major health facilities within the study area. Eight health centres in the Upper River Region provide primary health care and refer patients to Basse District Hospital when the need arises. Bansang Hospital serves the Central River Region and acts as the referral point for patients from Basse District Hospital. Both Health facilities also provide medical services to patients from the North Bank across The River Gambia as well as patients from Senegal.

### Study design

Verbal autopsies were conducted for deaths of children under-5 between September 01, 2019, and December 31, 2021. The VAs were coded by four physicians who worked in pairs. Each physician worked independently to assign a probable cause of death based on the information captured in the verbal autopsy forms. In each case, a primary and underlying cause of death was coded using information captured in the VA interview. The disease condition or terminal event which occurred just before the death was selected as the primary cause of death. The morbid condition or injury which initiated the chain of events leading to the death was selected as the underlying cause of death[29].

After coding, the results of both physicians were compared, and the points of disagreement were identified. These discrepancies were resolved by a consensus review of the verbal autopsy by the two physicians. The causes of death were assigned using a shortlist of deaths that could be ascertained from VA and mapped to the International Classification of Disease 10 codes[30].

### Data Collection

Trained field workers undertake household visits within the Basse and Fuladu West HDSS areas every 4 months. Data on all deaths, births, migrations, pregnancies, and the vaccination status of children are captured during visits. Verbal autopsies are conducted routinely when deaths are identified after a grieving period of about one month. The interviews are conducted using standardised closed-ended WHO VA questionnaires designed for neonates and children 1-59months of age[31]. Field workers were trained on each item of the questionnaire and how to conduct interviews with respect and understanding. During interviews, the questions are translated by field workers from English into the appropriate local language of the respondent. Corresponding data is captured into a tablet application using the Research Electronic Data Capture (REDCap) application database system[32].

### Statistical Analysis

All analyses were performed using Stata version 17 (StataCorp, College Station, TX, USA). The data were assessed for errors and missing values. Age was categorized into four groups to describe age-specific mortality (neonates 0-27days), 1-11 months, 12-23months, and 2-5 years). Variables with few numbers were grouped as “other”. Summary statistics were performed for key background characteristics such as sociodemographic factors, primary causes of death, underlying causes of death, and contextual factors (decision to seek care, access to medical care, and adequacy of care). The summary of the results is presented using frequency tables and percentages.

### Ethical Consideration

The collection of VA data is one of the procedures undertaken by the Basse and Fuladu West HDSSs. The HDSSs include hundreds of thousands of residents and constitute minimal risk observational research with data used by multiple research projects, non-government organisations, and government agencies. For these reasons, verbal consent is sought from the head of each household at each household visit. The operation and procedures of both the Basse and Fuladu West HDSS have been approved by the Gambia Government/MRCG Joint Ethics Committee (SCC number 1577).

## Results

### Socio-demographic characteristics

Verbal autopsies were conducted for 647 of 727 (89%) detected deaths of children aged less than 5 years during the observation period. Table 1 describes the socio-demographic characteristics of the evaluated deaths. Most of the deaths occurred in the neonatal period (37.1%, n=240). Outside of the neonatal period, 175 (27.1%) deaths occurred in the first year of life, 89 (13.8%) in the second year of life, and the remaining between 2 and less than 5 years of age (22.1%, n=143). There were comparable numbers of male and female deaths, with 324 (50.1%) females and 321 (49.6%) males.

**Table 1:**
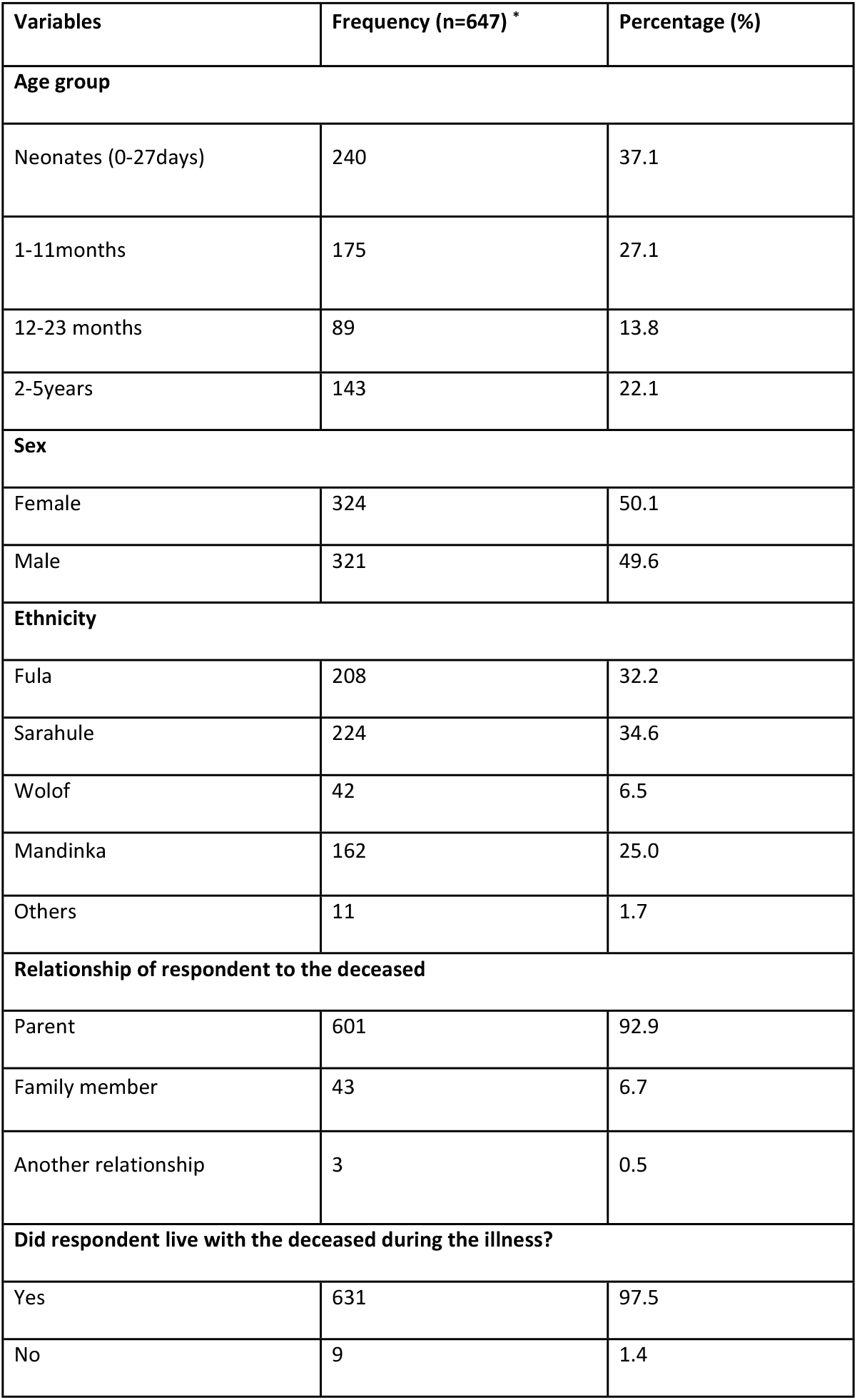
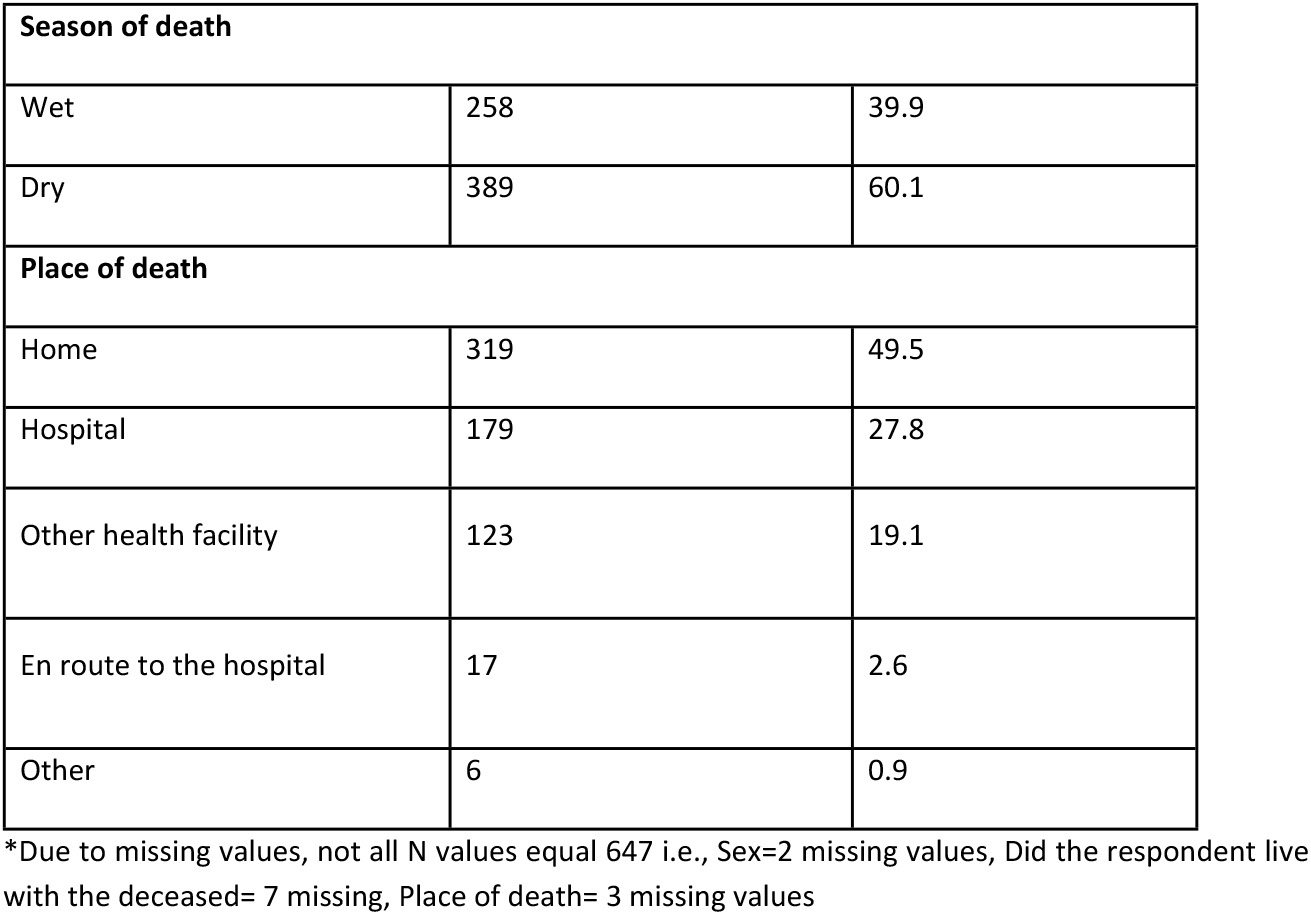
Socio-demographic characteristics of the deceased.

| Variables | Frequency (n=647) * | Percentage (%) |
| --- | --- | --- |
| **Age group** |  |  |
| Neonates (0-27days) | 240 | 37.1 |
| 1-11months | 175 | 27.1 |
| 12-23 months | 89 | 13.8 |
| 2-5years | 143 | 22.1 |
| **Sex** |  |  |
| Female | 324 | 50.1 |
| Male | 321 | 49.6 |
| **Ethnicity** |  |  |
| Fula | 208 | 32.2 |
| Sarahule | 224 | 34.6 |
| Wolof | 42 | 6.5 |
| Mandinka | 162 | 25.0 |
| Others | 11 | 1.7 |
| **Relationship of respondent to the deceased** |  |  |
| Parent | 601 | 92.9 |
| Family member | 43 | 6.7 |
| Another relationship | 3 | 0.5 |
| **Did respondent live with the deceased during the illness?** |  |  |
| Yes | 631 | 97.5 |
| No | 9 | 1.4 |

| Season of death |  |  |
| --- | --- | --- |
| Wet | 258 | 39.9 |
| Dry | 389 | 60.1 |
| Place of death |  |  |
| Home | 319 | 49.5 |
| Hospital | 179 | 27.8 |
| Other health facility | 123 | 19.1 |
| En route to the hospital | 17 | 2.6 |
| Other | 6 | 0.9 |

\*Due to missing values, not all N values equal 647 i.e., Sex=2 missing values, Did the respondent live with the deceased= 7 missing, Place of death= 3 missing values

Most of the deaths occurred amongst the Sarahule tribe (34.6%, n=224) and the Fula tribe (32.1%, n=208) which are the largest ethnic groups in the study area. Almost all the respondents who gave information relating to the circumstances of the deaths were family members, with 92.9% (n=601) being the parent(s) of the deceased. During the period of illness that led to death, 98% (n=631) of the respondents lived with the deceased. Concerning seasonality, 39.9 % (n=258) of the deaths occurred during the wet season, while the rest occurred during the dry season (60.1%, n=389). The majority of deaths occurred at home (49.5%, n=319) (Table 1).

### Contextual factors

In the final days of the illness, most parents knew that the deceased needed medical care (72.2%, n=464). However, in most cases, care was not sought outside the home (63.7%, n=421) and for those who sought any care outside the home, most of them went to a hospital or health centre (90.9%, n=210). Only a few respondents used a mobile phone to call for help during the period of illness (12.3%, n=79) (Table 2).

**Table 2:** Contextual factors.

| Contextual factor | Frequency (n=647) * | Percentage (%) |
| --- | --- | --- |
| **1. Decision to seek care** |  |  |
| **In the final days of illness, were there any doubts whether medical care was needed?** |  |  |
| Yes | 179 | 27.8 |
| **Sought care outside home** |  |  |
| Yes | 231 | 35.7 |
| **Where was care sought?** |  |  |
| Traditional Healer | 7 | 3.0 |
| Hospital | 57 | 24.7 |
| Health Centre | 153 | 66.2 |
| Others | 14 | 6.1 |
| **In the final days of illness, did anyone use a phone to call for help?** |  |  |
| Yes | 79 | 12.3 |
| **2. Access to medical care** |  |  |
| **Does it take more than 2hrs to get to the nearest hospital/health facility** |  |  |
| Yes | 76 | 11.8 |
| **In the final days before death, did s/he travel to a hospital or health facility?** |  |  |
| Yes | 460 | 71.3 |
| **3. Adequacy of medical care** |  |  |

| Did (s)he receive any treatment for the illness that led to death? |  |  |
| --- | --- | --- |
| Yes | 471 | 72.9 |
| Were there any problems with the way (s)he was treated in the hospital or health facility † |  |  |
| Yes | 8 | 1.7 |
| Were there any problems getting medications, or diagnostic tests in the hospital or health† facility? |  |  |
| Yes | 59 | 12.8 |

\*Due to missing data, not all N categories sum up to 647:
Missing values: In the final days of illness, were there any doubts whether medical care was needed? (4), Sought care outside home (2), In the final days of illness, did anyone use a phone to call for help? (2), Does it take more than 2hrs to get to the nearest hospital/health facility (2), In the final days before death, did s/he travel to a hospital or health facility? (2), Did (s)he receive any treatment for the illness that led to death? (1), Were there any problems with the way (s)he was treated in the hospital or health facility (1).
† Total frequency= 462

In most cases, there was the need to travel to a hospital or health facility (71.3%, n=460). In terms of access, most of the respondents lived less than 2hrs from a health facility (88.2%, n=569). In 72.9% (n=471) of cases, the deceased received some form of treatment during the illness For those who visited a health facility, there were no problems in how the deceased were treated (treatment, procedures, attitude, respect, dignity) in the hospital/health facility in most cases (98.3%, n=453). Only a few respondents reported having challenges getting medications or diagnostic tests at health facilities (12.8%, n=59) (Table 2).

### Primary causes of death

The primary causes of death are described in Table 3. Pneumonia (17%, n=110), diarrhoeal diseases (14.7%, n=95), sepsis (13.6%, n=88) and unspecified perinatal causes of death (11%, n=71) were the commonest primary causes of death. In the neonatal period, unspecified perinatal causes of death (29.6%, n=71), birth asphyxia (23.8%, n=57) and prematurity/low birth weight (17.1%, n=41) were the commonest causes of death. Outside the neonatal period, pneumonia (27.0%, n=110), diarrhoeal diseases (23.3%, n=95) and sepsis (21.6%, n=88) were the commonest primary causes of death. Of note, there were few malaria deaths (0.8%, n=5).

**Table 3.** Primary causes of death.

| Primary cause of death | Frequency (N=647) | Percentage (%) |
| --- | --- | --- |
| (0-59months) |  |  |
| Pneumonia | 110 | 17.0 |
| Diarrheal diseases | 95 | 14.7 |
| Sepsis | 88 | 13.6 |
| Other and unspecified perinatal cause of death | 71 | 11.0 |
| Birth Asphyxia | 57 | 8.8 |
| Prematurity Low Birth weight | 41 | 6.3 |
| Fresh stillbirth | 30 | 4.6 |
| Acute respiratory infection | 27 | 4.2 |
| Meningitis | 26 | 4.0 |
| Severe Anaemia | 26 | 4.0 |
| Cause of death unknown | 24 | 3.7 |
| Neonatal Sepsis | 16 | 2.5 |
| Road Traffic Accident | 6 | 0.9 |
| Malaria | 5 | 0.8 |
| Neonatal Pneumonia | 4 | 0.6 |
| Others | 21 | 3.2 |
| **Neonatal period (0-27days)** |  |  |
| Other and unspecified perinatal cause of death | 71 | 29.6 |
| Birth Asphyxia | 57 | 23.8 |
| Prematurity Low Birth weight | 41 | 17.1 |
| Fresh stillbirth | 30 | 12.5 |
| Neonatal Sepsis | 16 | 6.7 |
| Cause of death unknown | 13 | 5.4 |
| Severe Anaemia | 6 | 2.5 |
| Neonatal Pneumonia | 4 | 1.7 |
| Accidental fall | 1 | 0.4 |
| Macerated stillbirth | 1 | 0.4 |
| **1month-59months** |  |  |
| Pneumonia | 110 | 27.0 |
| Diarrheal diseases | 95 | 23.3 |
| Sepsis | 88 | 21.6 |
| Acute respiratory infection | 27 | 6.6 |
| Meningitis | 26 | 6.4 |
| Severe Anaemia | 20 | 4.9 |
| Cause of death unknown | 11 | 2.7 |
| Road Traffic Accident | 6 | 1.5 |
| Malaria | 5 | 1.2 |
| Severe Malnutrition | 4 | 1.0 |
| Contact with venomous animals and plants | 3 | 0.7 |
| Accidental exposure to smoke fire and flames | 2 | 0.5 |
| Assault | 2 | 0.5 |
| Other and unspecified cardiac disease | 2 | 0.5 |
| Accidental drowning and submersion | 1 | 0.3 |
| Others | 5 | 1.2 |

Fig 2 shows the primary causes of death grouped into infectious and non-infectious causes. Out of the 647 deaths, 57.3% (N=371) were due to infectious causes whiles the rest were due to non-infectious causes. Pneumonia, diarrhoeal diseases, and sepsis were the commonest infectious causes of death whereas birth asphyxia, prematurity/low birth weight, and fresh stillbirths were the commonest non-infectious causes of death.

**Figure 2.**
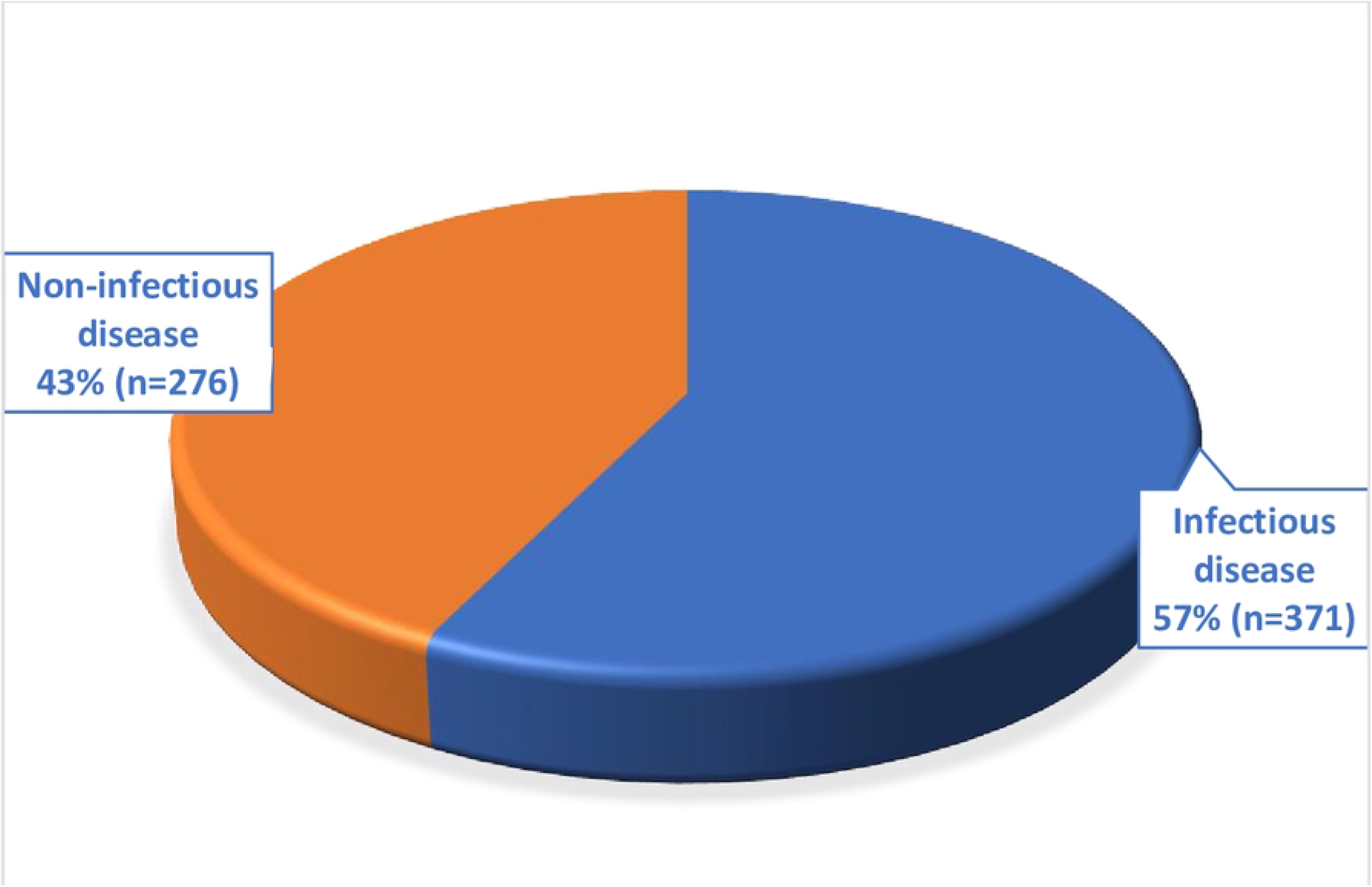
Primary causes of deaths grouped into infectious and non-infectious causes, 647 deaths.

### Underlying causes of death

Table 4 describes the underlying causes of death by age category. Severe malnutrition (28.6%, N=185), unspecified perinatal deaths (10.7%, N=69), pneumonia (10.2%, N=66) and prematurity/low birth weight (10.2%, N=66) were the commonest underlying causes of death.

**Table 4:**
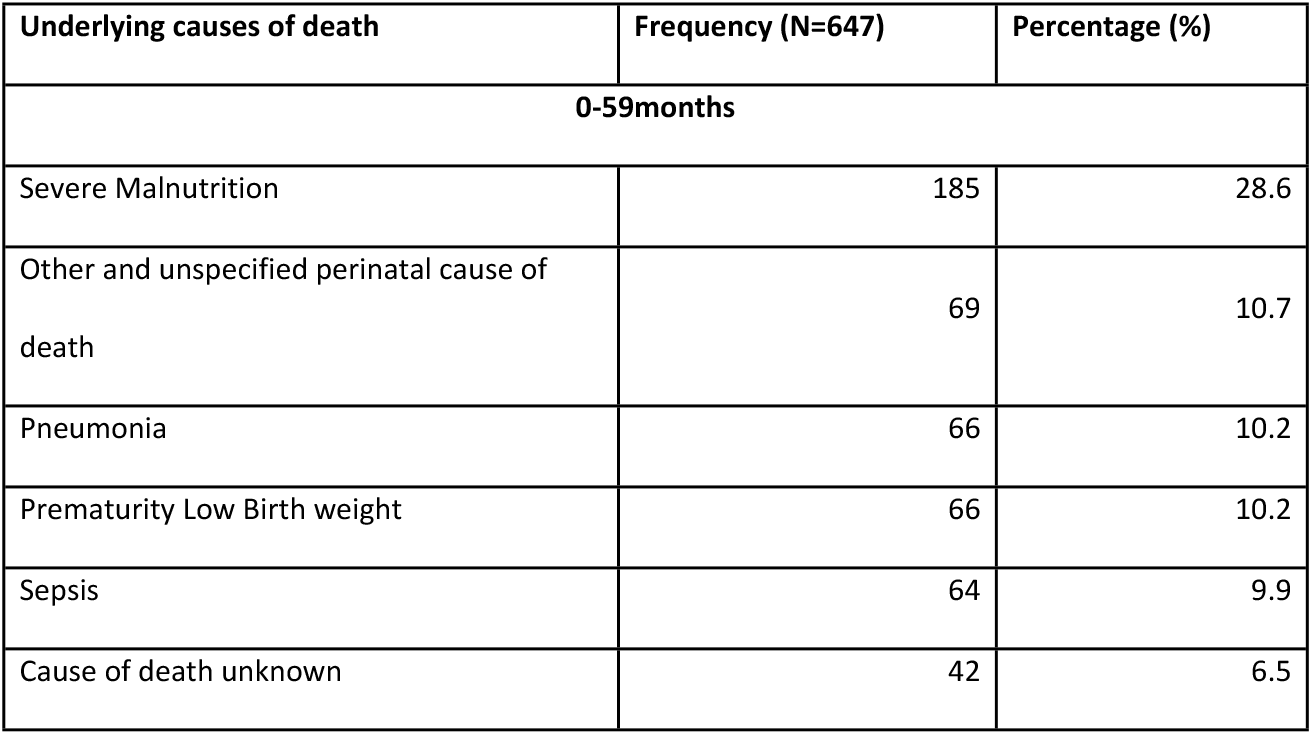
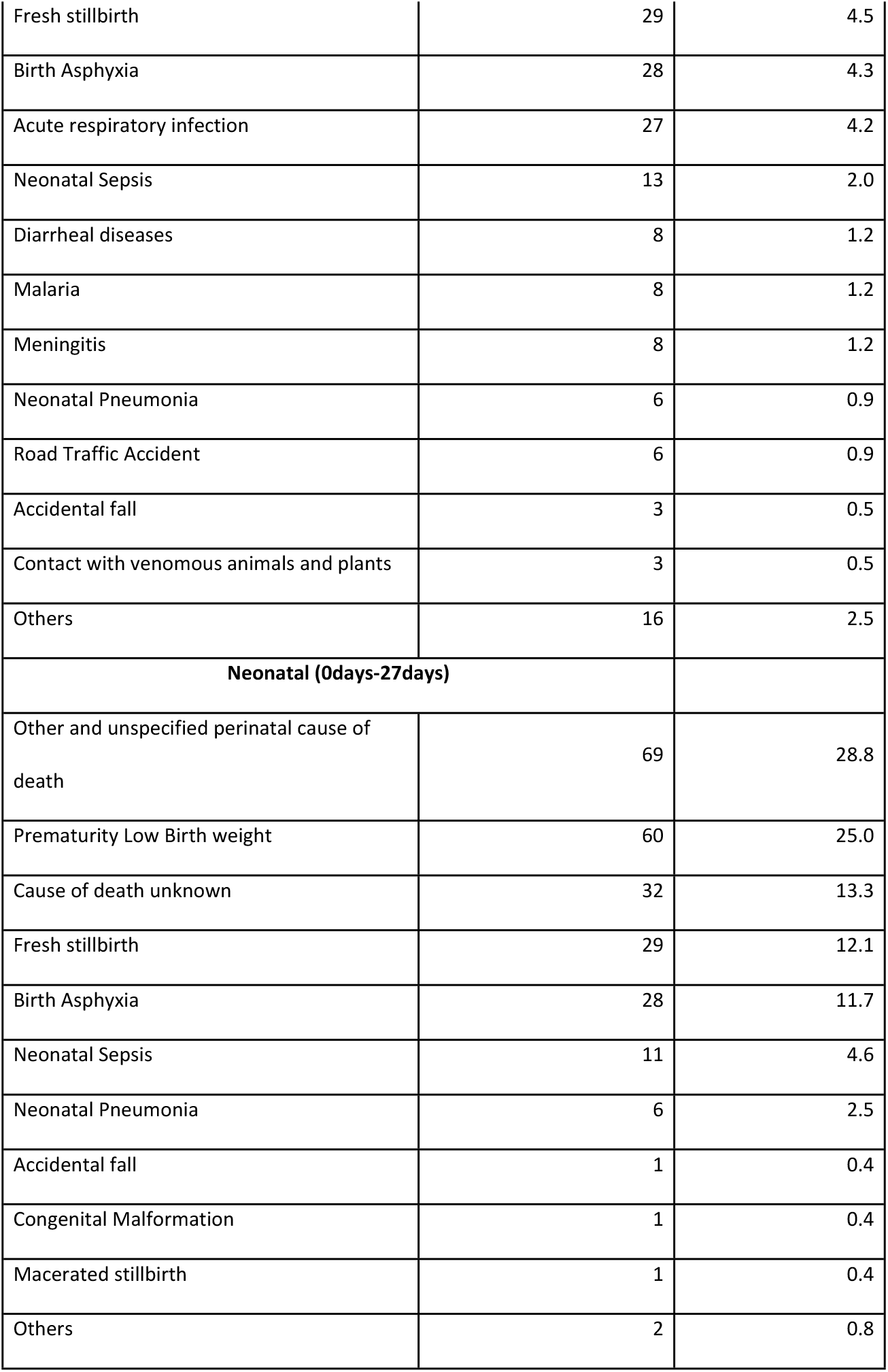
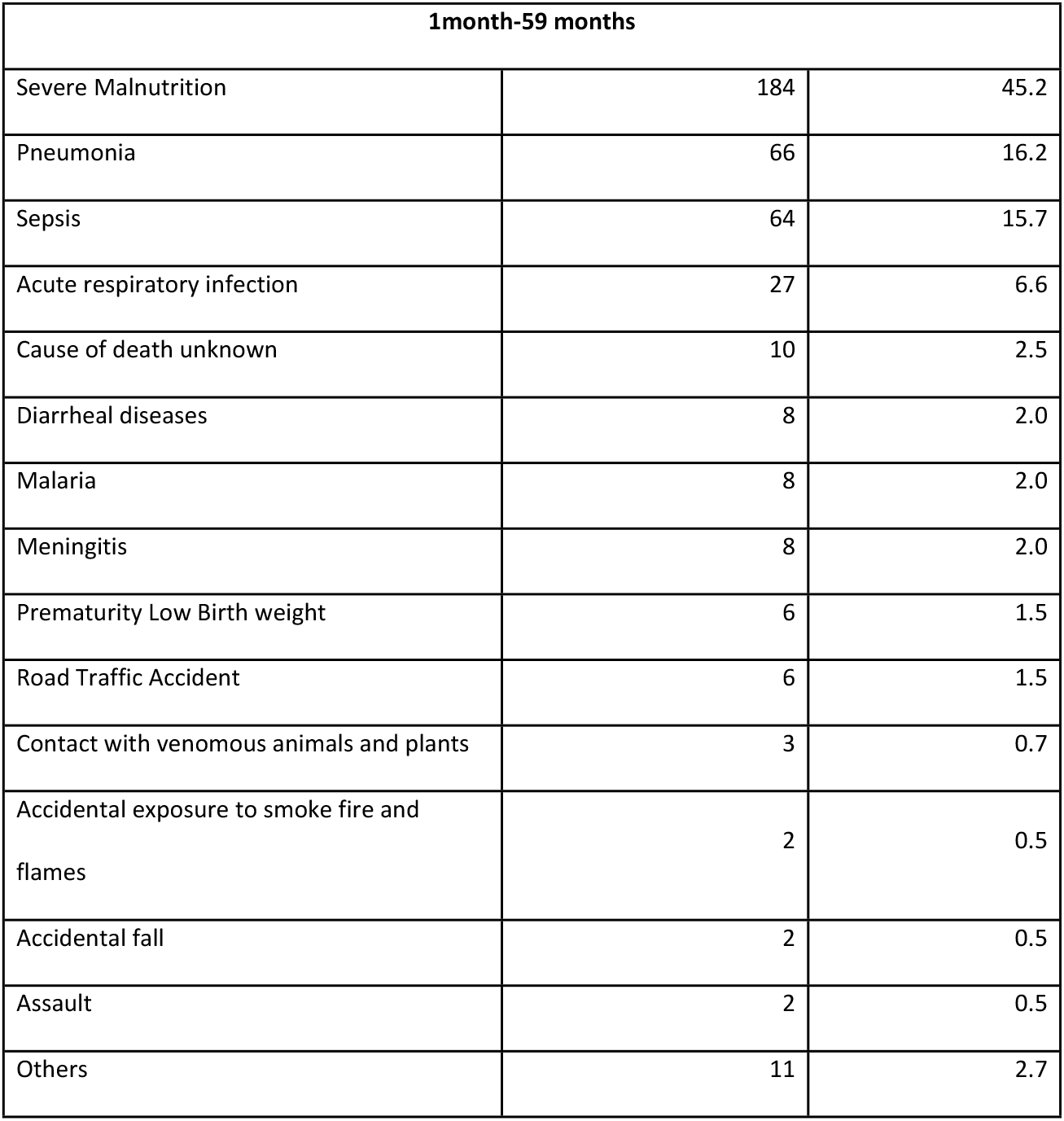
Underlying causes of death.

| Underlying causes of death | Frequency (N=647) | Percentage (%) |
| --- | --- | --- |
| **0-59months** |  |  |
| Severe Malnutrition | 185 | 28.6 |
| Other and unspecified perinatal cause of death | 69 | 10.7 |
| Pneumonia | 66 | 10.2 |
| Prematurity Low Birth weight | 66 | 10.2 |
| Sepsis | 64 | 9.9 |
| Cause of death unknown | 42 | 6.5 |
| Fresh stillbirth | 29 | 4.5 |
| Birth Asphyxia | 28 | 4.3 |
| Acute respiratory infection | 27 | 4.2 |
| Neonatal Sepsis | 13 | 2.0 |
| Diarrheal diseases | 8 | 1.2 |
| Malaria | 8 | 1.2 |
| Meningitis | 8 | 1.2 |
| Neonatal Pneumonia | 6 | 0.9 |
| Road Traffic Accident | 6 | 0.9 |
| Accidental fall | 3 | 0.5 |
| Contact with venomous animals and plants | 3 | 0.5 |
| Others | 16 | 2.5 |
| **Neonatal (0days-27days)** |  |  |
| Other and unspecified perinatal cause of death | 69 | 28.8 |
| Prematurity Low Birth weight | 60 | 25.0 |
| Cause of death unknown | 32 | 13.3 |
| Fresh stillbirth | 29 | 12.1 |
| Birth Asphyxia | 28 | 11.7 |
| Neonatal Sepsis | 11 | 4.6 |
| Neonatal Pneumonia | 6 | 2.5 |
| Accidental fall | 1 | 0.4 |
| Congenital Malformation | 1 | 0.4 |
| Macerated stillbirth | 1 | 0.4 |
| Others | 2 | 0.8 |

| 1month-59 months |  |  |
| --- | --- | --- |
| Severe Malnutrition | 184 | 45.2 |
| Pneumonia | 66 | 16.2 |
| Sepsis | 64 | 15.7 |
| Acute respiratory infection | 27 | 6.6 |
| Cause of death unknown | 10 | 2.5 |
| Diarrheal diseases | 8 | 2.0 |
| Malaria | 8 | 2.0 |
| Meningitis | 8 | 2.0 |
| Prematurity Low Birth weight | 6 | 1.5 |
| Road Traffic Accident | 6 | 1.5 |
| Contact with venomous animals and plants | 3 | 0.7 |
| Accidental exposure to smoke fire and flames | 2 | 0.5 |
| Accidental fall | 2 | 0.5 |
| Assault | 2 | 0.5 |
| Others | 11 | 2.7 |

Of the 240 neonatal deaths that were coded, 69 (28.8%) were unspecified neonatal deaths, 25 (25%) were due to prematurity/low birth weight and the cause of death was unknown in 32 (13.3%) cases. Amongst the deaths which occurred outside of the neonatal period, the trio of severe malnutrition (45.2%, N=184), pneumonia (16.2%, N=66), and sepsis (15.7%, N=64) were the top three underlying causes of death.

## Discussion

In this verbal autopsy analysis of deaths of children under-5 years of age in rural Gambia, we found that half of all deaths occurred at home. Pneumonia, diarrheal diseases, and sepsis were the leading causes of death. Over half of all primary causes of death were due to infectious diseases and severe malnutrition contributed to a third of the deaths. The highest number of deaths was in the neonatal period and the commonest causes of death in this age group were unspecified perinatal deaths, birth asphyxia, and prematurity/low birth weight. We also found that though most of the respondents were aware that the deceased needed medical care and lived within 2hrs of a health facility, care was not sought outside the home in most cases.

When deaths occur at home, they are not likely to be identified, recorded, or assigned a cause of death. This deprives decision makers of the opportunity to have complete data on disease burden among children under-5. One reason children may be dying at home instead of at health facilities is the reluctance of parents or caretakers to send them to the hospital when they are sick. This could be because they did not think the sickness was that serious or resorted to treatment at home. Another reason could be if treatment was sought in other places other than health facilities. However, in this study, only a few caretakers reported seeking care from traditional healers or pharmacies. Gupta N. et al. also found a similarly high number of home deaths in a matched case-control study in Rwanda[33]. With half of deaths occurring at home, it may be misleading to rely only on health facility data to characterise deaths of children under-5.

We also found that more than a third of deaths occurred in the neonatal period (37%). The high burden of deaths in the neonatal period was equally noted in an HDSS site in the northern part of India[34]. A significant proportion of births in The Gambia still occur without the supervision of skilled birth attendants[24] and may be a contributing factor to the unacceptably high number of neonatal deaths. From our experience, such neonates are often brought to the health facility a day or two after developing complications such as jaundice or sepsis.

Severe malnutrition and pneumonia were the commonest underlying and primary causes of death respectively. This is similar to an earlier study in Bangladesh where acute lower respiratory infection[35] was the most commonest cause of death. In that study, childhood wasting was additionally identified as the leading risk factor for lower respiratory infections. This is corroborated by a case-control study in Brazil which identified malnutrition as the most important risk factor for pneumonia in children[36]. These findings highlight the synergistic role acute malnutrition and pneumonia play in causing the demise of many children under-5. Apart from malnutrition, another risk factor that may have contributed to the high number of pneumonia deaths is the widespread use of charcoal and firewood for cooking in the study area. Mothers would usually have their children close by as they cook, exposing the children to smoke. This risk factor has been identified in a social autopsy as a major risk factor contributing to childhood deaths[37]. Therefore, measures aimed at reducing pneumonia deaths should also consider addressing malnutrition and indoor pollution from smoke.

Unspecified perinatal neonatal death was the commonest underlying cause of death in the neonatal period and the second overall underlying cause of death. This is the case because of the challenge in assigning specific causes of death using verbal autopsy for neonates who die in the first few days of life. This challenge was equally recognised by Baqui et al., who in a nationwide verbal autopsy analysis grouped neonatal deaths in the first three days of life as early perinatal deaths without further specifying[35]. A similar verbal autopsy study in Angola also identified unspecified neonatal deaths as the commonest cause of death in the neonatal period[38]. However, due to skip pattern errors in some of the neonatal verbal autopsy forms in our study, certain symptoms were not elicited. This further made it difficult to assign a specific cause of death in such cases.

One surprising finding in this study was the low number of deaths attributable to malaria since it is still a leading cause of death in many sub-Saharan countries. This observation however is due to the low prevalence of malaria in the The Gambia as a result of several years of malaria control interventions such as the use of insecticide-treated nets and mass drug administration[39-41]. Notwithstanding the non-specificity of malaria symptoms, there was a specific question in the VA questionnaire concerning whether the deceased had a recent positive malaria test. It is therefore not likely that malaria deaths were misclassified.

The social circumstances that contributed to the deaths were grouped using the three delays model (decision to seek care, access to care, and adequacy of care). Even though most of the caretakers did not doubt that the deceased needed medical care at the time of the illness, only a few of them sought care outside of the home. One reason could have been the issue of access to health facilities. However, the overwhelming majority reported that it did not take more than 2hrs to get to the nearest facility. A second reason for the reluctance in seeking medical care for the deceased could have been inadequate/quality of the care the children received at the health facilities. However, most of the respondents reported no problems in the way the children were treated, and in obtaining medications and other diagnostic tests.

Improved health-seeking behaviours through health education and promotion could contribute to reducing the number of under-5 deaths in rural Gambia. This, however, should be part of a comprehensive plan to improve health at all levels including more trained personnel and logistics at health facilities.

## Strengths/Limitations

A major strength of our study is using WHO’s standardised VA questionnaire. The advantage of this is that it allows our results to be compared to similar studies conducted using the same standard questionnaire. The relatively large sample size in this study also strengthens the conclusions drawn from the study. As noted by Uneke et al[20], the accuracy and reliability of verbal autopsies often depend on the type of healthcare system available and the experience of the physicians as far as local disease aetiology and epidemiology are concerned. The four physicians who partook in analysing the VA interviews and assigning causes of death were trained on the VA tool before the coding began. All of them have worked in the study area and therefore understand the local disease epidemiology and common presentations at the health facilities.

Another strength of this study is that most of the respondents were first-degree relatives of the deceased, so more accurate information was likely to be collected by the field workers. Almost all the respondents lived with the deceased during the period of illness, further enhancing the credibility of the information they provided during the VA interviews.

However, verbal autopsy analyses have some inherent limitations. Translation of the VA questionnaire from English to the local languages may have caused some meaning to be lost or misinterpreted. To mitigate this, only field workers who were fluent in the language and understood the culture and customs of the respondent were allowed to conduct such VAs. The field workers were also trained on the verbal autopsy tool and were conversant with the use of the application for collecting the VA data.

Possible recall bias is another well-known limitation of verbal autopsies. To mitigate this in our study, we ensured that VAs were conducted as soon as possible, being mindful however to allow at least a month for relatives to mourn the deceased. Though all attempts were made to conduct all pending VAs, we were unable to perform the VAs for 11% of the deaths mainly due to the migration of primary respondents outside the study area. As indicated earlier, specific deaths could not be assigned to some neonatal deaths due to inadequate signs and symptoms in the verbal autopsy forms. Such deaths were classified as unspecified perinatal deaths.

## Conclusion

Verbal autopsies are useful in bridging the current gap in countries with weak systems for detecting and assigning childhood causes of death. According to VA analysis, half of deaths amongst children under-5 in rural Gambia occur at home. Over half of deaths are due to infectious diseases. Pneumonia and severe malnutrition are the commonest primary and underlying causes of death respectively. Though most respondents were aware of the need for medical care and lived close to health facilities, most did not seek medical care. Improved health-seeking behaviour may reduce childhood deaths in rural Gambia.

## Data Availability

All relevant data are within the manuscript and its Supporting Information files

## Supporting Information

S1 File. Minimal Data Set V 1.0

S2 File. VA Coding list_V1.0.pdf

S3 File. WHO_VA_2016_v1.5.3.XLS_form_for_ODK

## Acknowledgements

We thank all the grieving families who agreed to be part of this study. We also thank all the hardworking field workers who undertook the verbal autopsy interviews. We are grateful to the HDSS team at the Medical Research Council Unit The Gambia at London School of Hygiene and Tropical Medicine for overseeing and supervising the collection of verbal autopsies.

